# How floods may affect the spatial spread of respiratory pathogens: the case of Emilia-Romagna, Italy in May 2023

**DOI:** 10.1101/2024.09.20.24314056

**Authors:** Claudio Ascione, Eugenio Valdano

## Abstract

The negative impact of floods on public health has been increasing, as climate change makes these events more frequent and intense. Floods are known to cause direct injury and favor the spread of many waterborne and vector-borne pathogens. Their effect on the circulation of respiratory pathogens, like influenza and SARS-CoV-2, is, however, still unclear. In this study, we quantify this effect through the analysis of large-scale behavioral data coupled to mathematical models of epidemic spread. We focus on the devastating floods occurred in Italy in 2023 and measure how they impacted human contact patterns within and between communities. We find a substantial increase in contacts occurring 3 weeks after the floods, both among residents of the affected areas and between them and those living in distant, unaffected areas of Italy. Then, through mathematical simulations, we determine that these disrupted contact patterns can carry a circulating pathogen to previously unaffected geographic areas, as well as increasing infection counts across the country. Our findings may help set up protocols to use large-scale human contact data to contain epidemic outbreaks before, during and in the aftermath of floods.

## Introduction

Climate change is increasing the frequency and intensity of floods [1–3]. This poses a clear threat to public health, as floods increase the risk of direct injury [4], hamper access to healthcare [5] and facilitate the emergence and circulation of infectious disease epidemics. In this context, floods are known to increase the spread of pathogens that are transmitted through environmental exposure[6], like those causing cholera [7], other diarrheal diseases [8], skin and soft-tissue infections [9], Legionnaires’ disease [10]. They also increase the risk of zoonoses, like leptospirosis [11], and mosquito-borne diseases, like Chikunguya [12]. The effect of floods on the epidemics of directly-transmitted respiratory pathogens, like influenza viruses and coronaviruses (notably SARS-CoV-2) is, instead, less known. Here, we posit that the changes in human contact patterns that floods bring about may have a considerable effect on the circulation of respiratory pathogens, and aim to quantify this effect.

Human contact patterns are heavily disrupted during, and in the aftermath of, extreme climatic events [13–16]. In the case of floods, people may be confined at home in the beginning, and then relocated to shelters [17]. At the same time, aid may need to flow in from unaffected areas. These kinds of changes in the patterns of close-proximity interactions and time spent indoors are known, from other contexts, to influence the spread of respiratory diseases [18–24], and their effects are being studied also in the contexts of other extreme climatic events, like wildfires [25].

In this study, we aimed to characterize the potential link between floods and epidemic of respiratory pathogens mediated by changes in human contact patterns. To this end, we focused on the floods occurred in the Italian region of *Emilia-Romagna* in May, 2023 (see Fig.1). 12,241 hectares of urban and agricultural land as well as roads and infrastructure were flooded for five weeks after anomaloulsy intense rain [26]. This caused the displacement of around 30, 000 people and an estimated damage of C10 billion [27, 28]. We reconstructed and analyzed human contact patterns before, during and in the aftermath of the event using data provided by Meta’s Data for Good Lab [29]. Then, we used a mathematical model to simulate the spread of a COVID-19-like epidemic and estimate the effect that the flood had on its spread by means of the changes in contact patterns.

**Fig 1.**
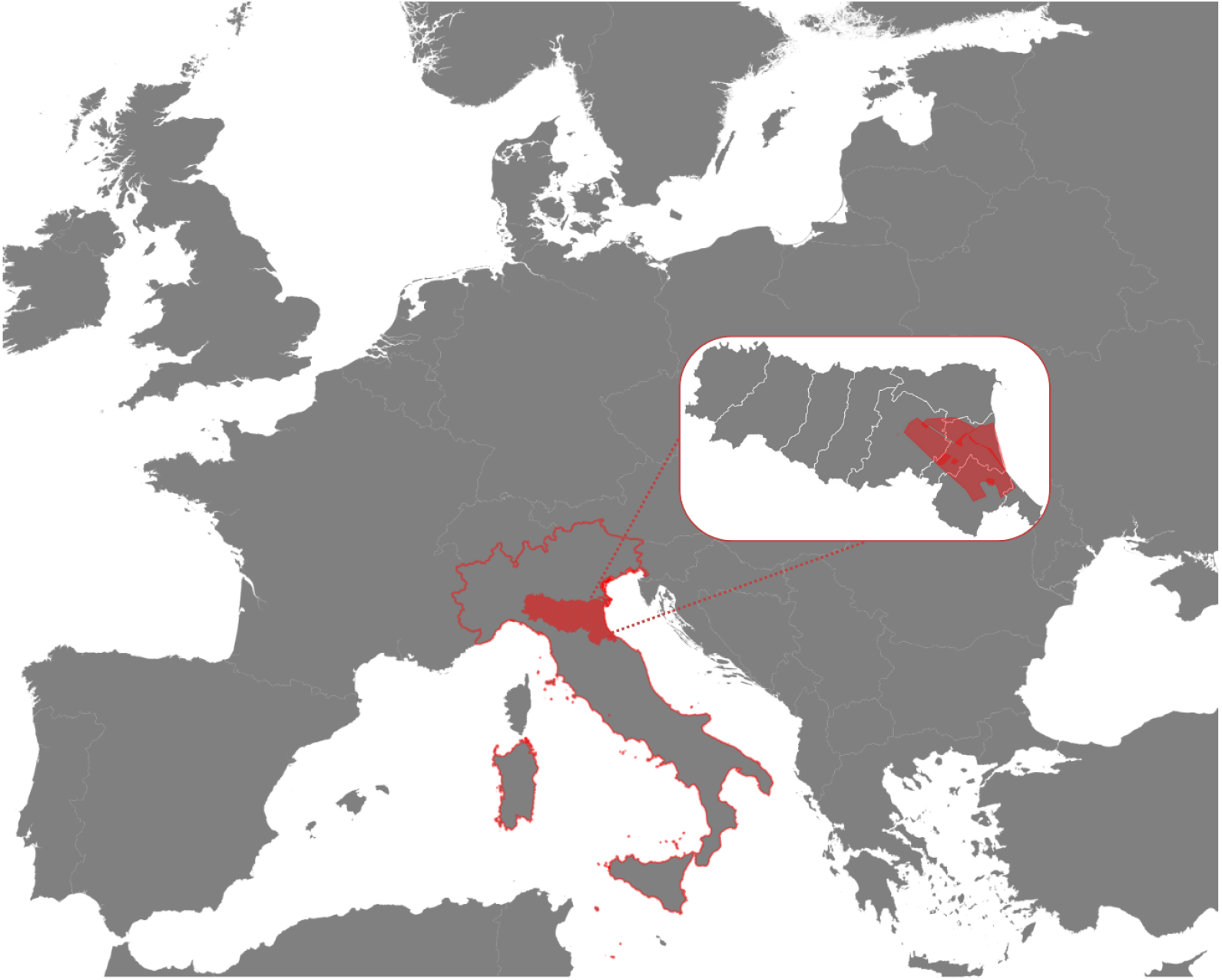
Location of Italy in Europe (red outline), Emilia-Romagna in Italy (red), the areas affected by the flood in May 2023 according to [26] (red in inset) overlaid on the provinces of Emilia-Romagna (white outline in inset).

Our findings may help step up public health preparedness for a type of compound emergency – floods – that climate change is making graver and more common.

## Data and Methods

### Timeline of the Floods in Emilia-Romagna

The major flood in Emilia-Romagna, Italy started on May 16th. We set a weekly time scale to match that of the contact data and set the week from May 8th to May 15th to be *week 0*, and counted time relative to it. This means that the flood started between week 0 and week 1. In the analysis of contact patterns we specified one time point using this unit and resolution, i.e., the week of the measured contacts relative to week 0. Also, for epidemic simulations, we specified the start time – the week, relative to week 0, when the outbreak started – and the observation time – the week, relative to week 0, when we measured the epidemic size. Also, we defined a *baseline period* from week −3 to week 0 included, which we used to compute relative variations in contacts attributable to the flood and to design the counterfactual scenario for the simulated epidemics.

### Data

#### Flood

We gathered data from the ESA Copernicus Database, which is a European program for global satellite monitoring. One of its features is emergency management, in which they study the effect of critical climatic events on the local impacted areas, quantifying it in terms of land coverage, economic loss and acres affected. For our purposes, we used data about the Emilia-Romagna Floods of May 2023 to determine the geographical areas impacted by the climatic event [26].

#### Contacts

Human close-proximity contact data was provided by Meta Data for Good Database [29] in the form of Colocation maps. Colocation maps measure the rate at which two individuals, residing one in spatial community *x* and the other in spatial community *y*, are found themselves in close proximity. They have a weekly time resolution and, in Italy, the spatial resolution of *provinces*, level-2 administrative divisions broadly matching the European Union standard *NUTS-3*. Details on how Colocation maps are built from location data from mobile devices can be found in [29]. For contacts occurring among residents of the same province, colocation maps are stratified on whether contacts occurred when two people were both the level-16 Bing tile where they were estimated to reside, or not. We could then discriminate between contacts attributable to be occurring between members of the same household or in the community. We referred to household contacts and community contacts, respectively. For our purposes, we selected data from Italy across 33 weeks from April 17th, 2023 to December 4th, 2023. This gave 33 Colocation maps for household contacts and 33 Colocation maps for community contacts, at a weekly resolution.

We also used social contact rates from the *SOCRATES* project, a broadly used source to inform epidemic modeling, stratified by location (household, community) [30, 31].

## Methods

### Internal and external contact rate

To analyze contacts among residents of the same province and among residents of different provinces, we defined three metrics: household contacts, community contacts and external contacts. Let 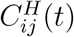 be the within-household and 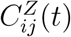 the outside-household Colocation map between community *i* and community *j* at time *t*. These quantities inherently define two matrices: the within-household contact rate matrix *C*^*H*^ ∈ ℝ ^*n,n*^ and the outside-household contact rate matrix *C*^*Z*^ ∈ ℝ ^*n,n*^, with *n* indicating the number of provinces. Note that 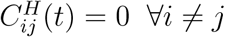 as people cannot reside in the same household but in different provinces. Let then *N*_*i*_ be the resident population of province *i*, obtained from Ref. [32]. Then, following the methodology described in Ref. [29], 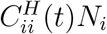 measures the household contact rate in *i*, 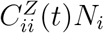 the community contact rate *i* and 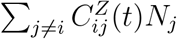 the external contact rate of *j*, i.e., the rate at which residents of *i* establish contacts with residents of other provinces. Denoting 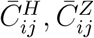 the Colocation maps average over the baseline period, we then could compute the relative variation of each contact rate, at different times, with respect to the baseline.

#### Epidemic model

To simulate the population-level spread of an epidemic of a directly-transmitted respiratory pathogen we used a stochastic metapopulation model. We assumed a SusceptibleExposed-Infected-Removed compartmental model to describe disease natural history[33–36], whereby infectious individuals can transmit the disease to susceptible ones, which become exposed (infected but not yet infectious). At predetermined rates, exposed individuals become infectious and infectious individuals become removed, i.e., are no longer infectious and cannot be re-infected.

At initialization, all individuals in all provinces are in the susceptible compartment. If *S*_*i*_ measures the number of susceptible individuals in province *i* then *S*_*i*_ = *N*_*i*_ at the start. We then initialized the outbreak by setting one infectious individual in the province where the outbreak had origin. We selected the province of Rimini as it was the most strongly affected by the flood (for alternative choices, see Supplemental Material). If *I*_*i*_ is the number of infectious individuals in province *i* then, at the start, *I*_*Rimini*_ = 1, *S*_*Rimini*_ = *N*_*Rimini*_ − 1, *I*_*i*_ = 0, *S*_*i*_ = *N*_*i*_ ∀*i*≠ *Rimini* at the start. Analogously we defined *E*_*i*_, *R*_*i*_ as the number of individuals in the exposed and removed compartment. All these quantities change along the simulation but we assumed the resident population to be constant: *S*_*i*_(*t*) + *E*_*i*_(*t*) + *I*_*i*_(*t*) + *R*_*i*_(*t*) = *N*_*i*_.

The metapopulation model is a stochastic process, defined as such:

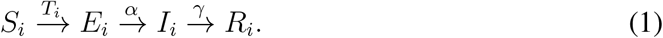

*T*_*i*_, *α, γ* are the transition rates, measured in days^*−*1^. As customary[37], to simulate it, we discretized the process using a small discretization window Δ*t* = 1 day. Therefore the individual probability of transition from E to I during a discretization time step is *P*_*α*_ = 1 − *e*^*−α*Δ*t*^ and the probability of recovery (I to R) is *P*_*γ*_ = 1 − *e*^*−γ*Δ*t*^. The individual probability of becoming infected instead depends on the force-of-infection at a specific time, which is a combination of the possibility of being infected by a member of the same community, or another community, mediated by the contact rates. This can be written using an effective Colocation map 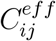 so that the transmission rate in the stochastic process reads as follows (see Ref. [29]):

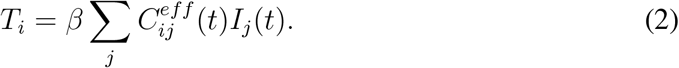

To determine *C*^*eff*^, we noted that Colocation Maps, by construction, are effective at measuring variations in contact rates but lack a transmission-specific definition of what contact are. To overcome this, we rescaled the average Colocation maps values during the baseline period using SOCRATES social contact data. Specifically, we assumed that the number of household contacts during the baseline period, which is proportional to 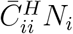, equals the number of household social contacts from SOCRATES in each community. Similarly, we assumed that community contacts, which are proportional to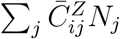, equals the number of community social contacts from SOCRATES. Finally, we encoded the facts that community contacts are associated with a lower transmission rate than household contacts through a damping parameter *ω* = 0.3 [38, 39]. This completely determines the form of 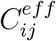 that appears in Eq. 2:

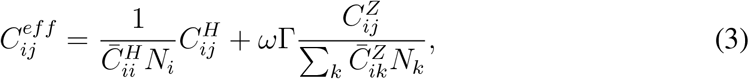

where Γ is the ratio between the number of community contacts over household contacts reported by SOCRATES. More details of the derivation of Eq. (3) are in Supplemental Material. With the individual transition rates, we simulated the total number of transitions per discretization step by sampling from binomial distributions: in community *i*, the number of transitions from E to I is binomially distributed with *n* = *E*_*i*_ and *p* = *P*_*α*_, the number of transitions from I to R has *n* = *I*_*i*_ and *p* = *P*_*γ*_ and finally the number of incident infections *n* = *S*_*i*_ and *p* = *T*_*i*_. We fixed the rates *α, γ* from the literature, assuming a COVID-19-like disease [40]. This left *β* as the only free parameter of the model, which we used to tune the basic reproductive ratio of the emerging outbreak, via customary relationship

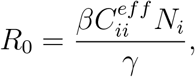

where *i* is the community in which the outbreak starts [40, 41]. In the simulations, at each time step, we computed the total attack rate for each province *i*: i.e. *AR*_*i*_(*t*) = (*E*_*i*_(*t*) + *I*_*i*_(*t*) + *R*_*i*_(*t*)) */N*_*i*_ and performed statistical analysis on this quantity as a proxy for the state of the epidemic at each instant *t*. For each scenario tested, we performed 20, 000 independent simulation runs. Then, we computed the median (50^*th*^ percentile) attack rate value to estimate the median epidemic outbreak size and the 99.9^*th*^ percentile to estimate the size of large-scale epidemic outbreaks. To estimate the geographic shape of epidemics we estimated the median attack rate in each province and the probability that a province was reached by the epidemic.

We simulated different epidemic scenarios, by varying the basic reproduction ratio in the range [0.5, 2.5], the origin of the outbreak, the week when it started (*outbreak week* henceforth) and in which week the epidemic outcome was observed. To estimate the impact of the flood, we measured changes in outbreak size and shape with respect to a counterfactual scenario in which we replaced observed Colocation maps in weeks 0 through 3 with the baseline. Then, we estimated relative changes in the aforementioned quantities by bootstrapping 5, 000 runs from the scenarios under study and the counterfactual.

## Results

### Contact patterns

We analyzed contacts at week 0 (week right before the event), week 1 (immediate aftermath), week 3 (after the event) – see Fig. 2.

**Fig 2.**
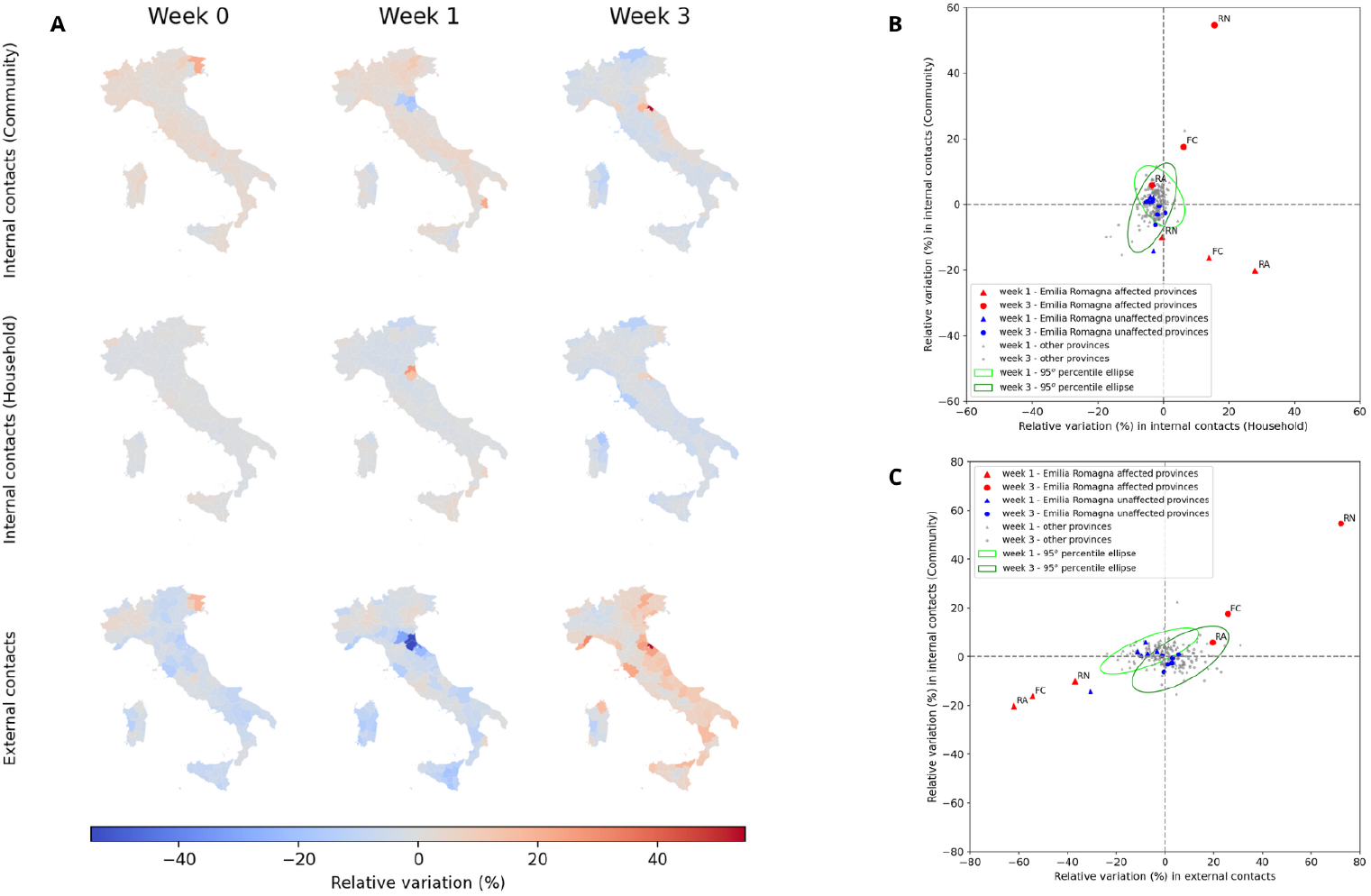
**A**. Administrative level 2 maps (province) of Italy, colored according to the relative variation value of the community contacts, household contacts and external contacts with respect to the baseline. For each type of contact, we observed the values in 3 specific weeks (Week 0 is the week May 8th-14th, Week 1 May 15th-21st, Week 3 May 29th-June 6th) **B**. Scatterplot of the relative variation of the household vs the within-community contacts. The triangles week 1, the circles week 3. In red, we highlighted the affected provinces (Rimini, Ravenna, Forlì-Cesena); in blue, the unaffected ones in the same region of the affected ones (Bologna, Parma, Piacenza, Reggio-Emilia, Modena, Ferrara). The ellipse in light green is the 95*th* percentile ellipse built on the distribution of all provinces at week 1, the one in dark green is the same ellipse built on the distribution at week 3. **C**. Scatterplot of the relative variation of the external and withincommunity contacts. Symbol and color codes are the same as **B**.

At week 0 there was no visible change in contacts with respect to baseline, across contact types (see Fig. 2**A**): variations in household contacts ranged from −3% to +4%, in community contacts from −6% to +22% and in external contact from −14% to +18%. The only exception was the province of Udine in northeastern Italy bordering Slovenia, far from the flood affected area, which showed a moderate increase and likely attributable to a mass gathering (Adunata Nazionale degli Alpini [42]).

At week 1, household contacts had a marked increase from baseline in the affected provinces, peaking at +28% in Ravenna. Instead, community contacts and external contacts decreased in affected provinces. This decrease was most marked for external contacts, with −37% in Rimini, with −62% in Ravenna. In addition, external contacts decreased also in provinces which were not directly affected by the flood but bordered affected provinces, like Bologna (−31%). At week 3, household contacts were still moderately higher than baseline in affected provinces (+16% in Rimini), community contacts were however markedly higher than the baseline and external contacts also increased above the baseline, especially in Rimini (+72%). A moderate increase in external contacts was recorded in week 3 across Italy, likely to be attributable to other factors, but the increase recorded in the affected provinces was substantially higher (see Fig. 2**C**, except for Ravenna). More broadly, Fig. 2**B,C** compares variations in contact metrics in the affected provinces and compare them to variations in unaffected provinces. Except for Ravenna, all variations are consistently outside the 95*th* percentile computed on all provinces. Notably, the anomalous behavior did not consistently extend to unaffected provinces close to affected areas.

Fig. 2**B** also shows the correlation in the disruption of household and community contacts: in week 1, the increase in household contacts was associated with a decrease in community contacts, while in week 3 they both increased with respect to the baseline. Analogously, Fig. 2**C** shows that community and external contacts decreased (week 1) and then increased (week 3) together.

### Simulated epidemic outbreaks

We present here results for epidemics starting in the province of Rimini, in weeks −2, 0, +2 and observed four weeks after the start of the epidemic. Other choices are presented in the Supplemental Material. First, we examined the impact of the flood on the size of the epidemic. Figure 3 shows the variation in median epidemic size and in the size of large epidemics, inside and outside the starting province of Rimini, for varying basic reproductive ratio (*R*_0_) and outbreak week. Overall, epidemic sizes either increased or remained the same with respect to the counterfactual scenario. The size of epidemics inside Rimini, both median and large, had the same qualitative behavior as a function of *R*_0_ across outbreak weeks: the effect of the flood increased up to around the epidemic threshold (*R*_0_ = 1), stayed constant and then dropped at *R*_0_ = 1.5 − 1.75. In magnitude, the effect was stronger for outbreak week +2 (an epidemic starting two weeks after week 0): median epidemic size was, for the *R*_0_ = [1 … 1.5] plateau, 20% larger than in the counterfactual scenario and large epidemics were 10% larger (see Fig. 3**E,F**). The size of the epidemics outside the starting province of Rimini followed a similar trend, with two exceptions. First, the flood-attributable increase in epidemic size was marked even for large values of *R*_0_, with the plateau extending to almost *R*_0_ = 2.5. (see Fig. 3**A,B,C,D**). Second, the increase in median size when the epidemic started in week 2 (see Fig. 3**E**) was substantially below the epidemic threshold, surpassing +30% for *R*_0_ = 0.5.

**Fig 3.**
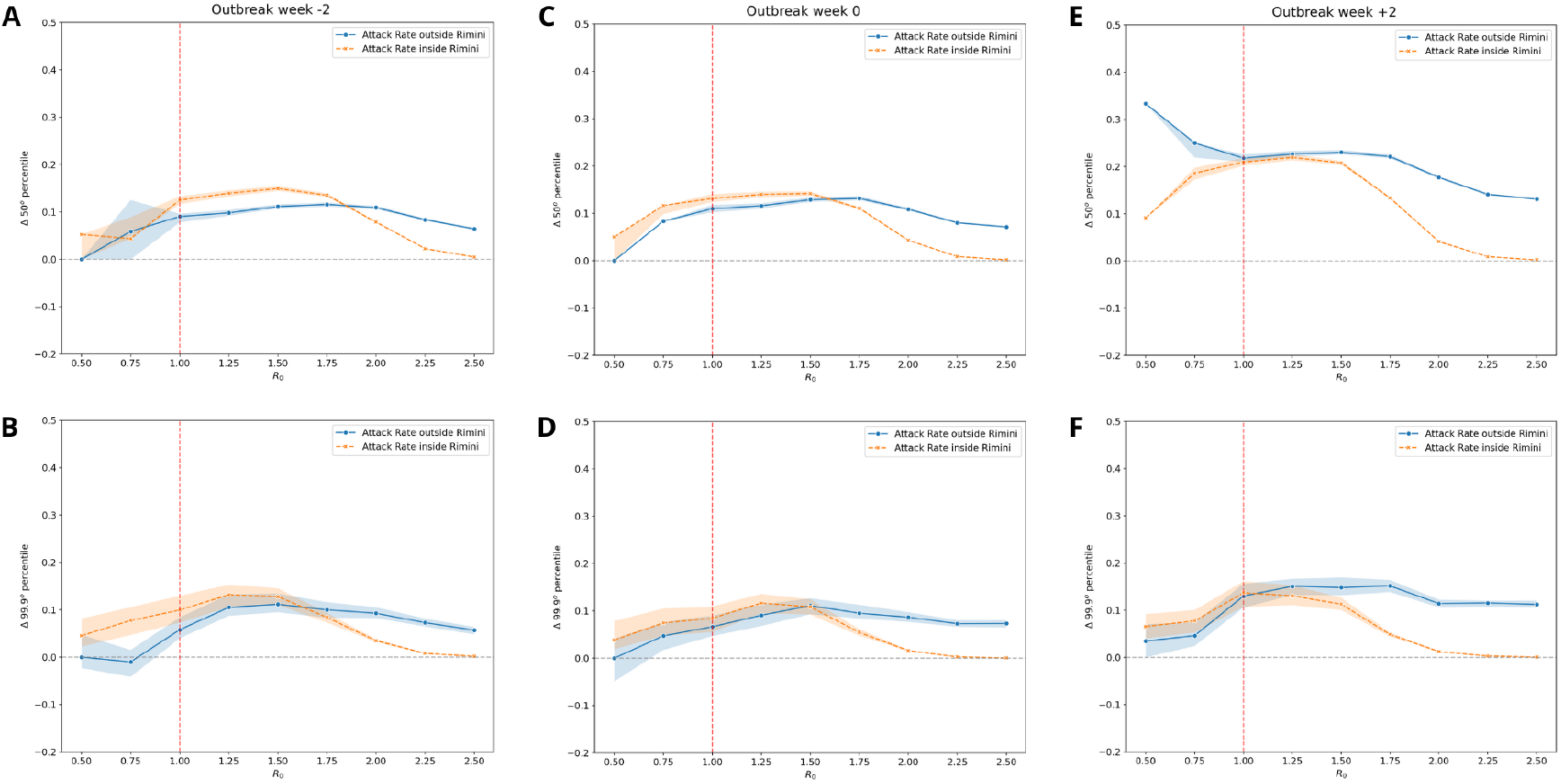
Relative variation with respect to the counterfactual scenario of the median (**A,C,E**) and the 99.9^*th*^ percentile (**B,D,F**) value of the Attack Rate outside and inside the province of Rimini for different cases of outbreak week at varying values of the local reproduction ratio. In orange, the Attack Rate inside the province of Rimini. In blue, the Attack Rate outside the province of Rimini. The red dotted line reports the epidemic threshold (*R*_0_ = 1). The shaded areas report the 95%.

To measure the impact of the flood on the spatial spread of simulated epidemics in Italy, we computed for each province either the risk that that province be reached by the epidemic or the relative median size of the outbreak there, both relative to the counterfactual. We focused on the impact of the flood on the probability that an epidemic well below the epidemic threshold (*R*_0_ = 0.5) reaches a province (Fig. 4**A,C,E**) and the impact on the local median epidemic size while well above the threshold (*R*_0_ = 2.5 - Fig. 4**B,D,F**). In the former, for outbreak weeks −2, 0 (see Fig. 4**A,C**) the impact was highly province-dependent, with the flood causing some provinces to become more likely to be reached, other less likely. Notably, effects are marked for provinces near the affected area as well as far from it. For outbreak week +2 (see Fig. 4**E**), the flood instead caused an increase in the probability to be reached in almost all the provinces of Italy. For the above-threshold epidemic, outbreaks weeks −2, 0 (see Fig. 4**B,D**) exhibited a homogeneous increase in local outbreak size, around +10% for all the provinces. For outbreak week +2 (see Fig. 4**F**) the picture is instead heterogeneous, with some provinces seeing a relative increase in outbreak size of up to +40%, while some, albeit very few, exhibiting a slight decrease.

**Fig 4.**
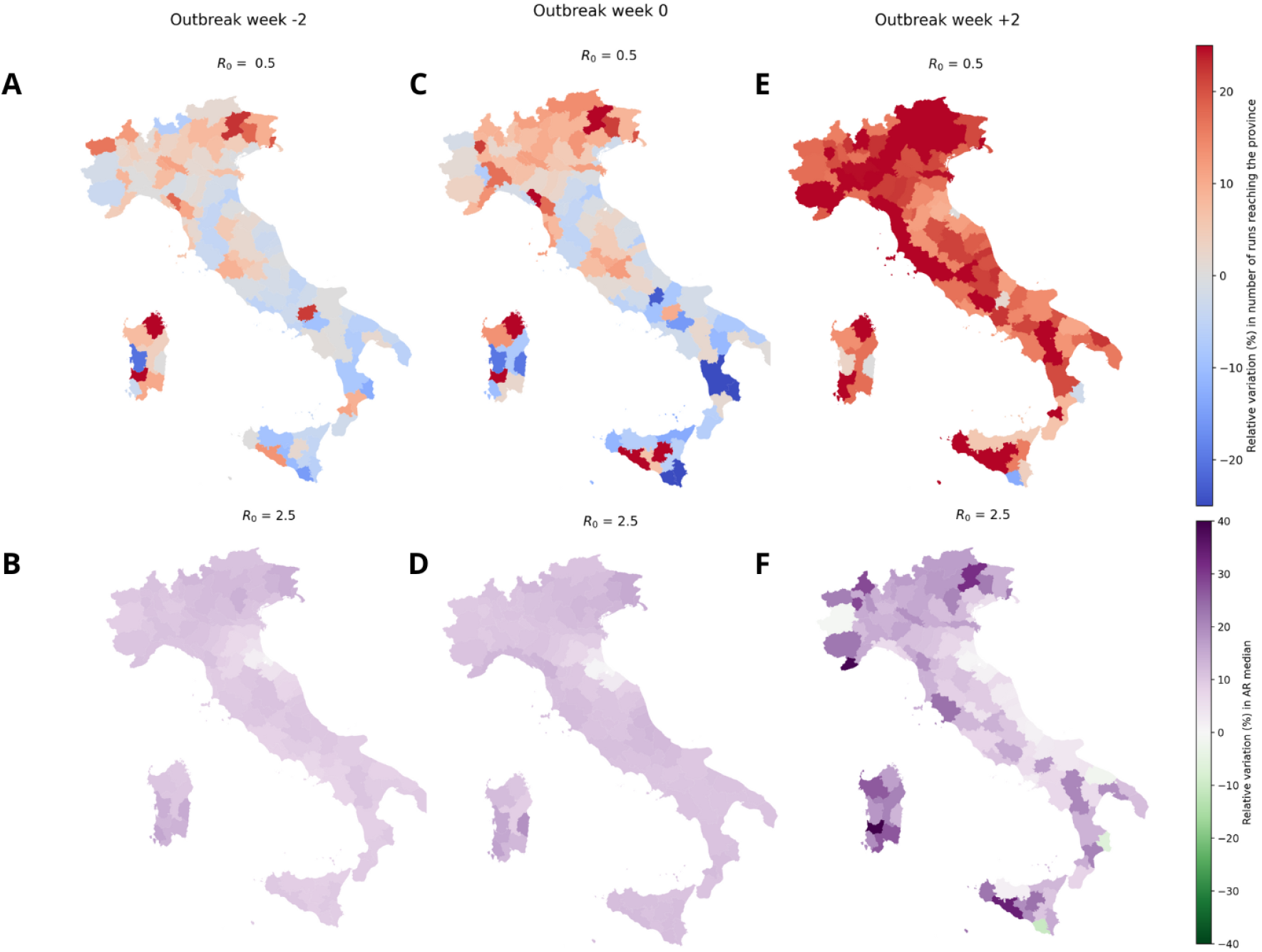
**A,C,E**. Administrative level 2 maps (province) of Italy, colored according to the relative variation (%) in the probability of the epidemic reaching a province, for 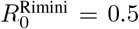, at varying outbreak week, with respect to the counterfactual case. **B,D,F**. Administrative level 2 maps (province) of Italy, colored according to the relative variation (%) in the median local Attack Rate, for 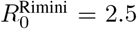, at varying outbreak week, with respect to the counterfactual case.

Finally, we investigated to which extent these spatial heterogeneities in the way the flood affected the simulated epidemics could be directly attributable to the direct changes in the contact network between provinces that the flood induced. Figure 5 investigates the correlation between the metrics of Fig. 4 and the relative change in the contact rate between each province and the province where the simulated epidemics started (Rimini). For example, let us consider *R*_0_ = 0.5, outbreak week +2 and the relative change in the probability that each province is reached by the epidemic (see Fig. 5**E**). We measured the extent to which the change in probability in province *j* was correlated with the in the contact rate between Rimini and *j* in week 3 (the week with stronger external changes, see Fig. 2), relative to the baseline. This correlation was strong, with a 0.78 (*pv* = 0.00) Pearson’s correlation coefficient. Overall the correlation was strong and significant for the scenario below the epidemic threshold (*R*_0_ = 0.5) across outbreak weeks (see also Fig. 5**A,C,E**), with correlation coefficient 0.77 for outbreak week −2 and 0.80 for outbreak week 0. In the above-threshold scenario (*R*_0_ = 2.5) the correlation was markedly weaker but still significant (*pv <* 0.05) for outbreak week −2 (correlation 0.58) and outbreak week 0 (correlation 0.51). There was instead no significant correlation in outbreak week +2 (correlation 0.15, *pv* = 0.12) (see also Fig. 5**B,D,F**).

**Fig 5.**
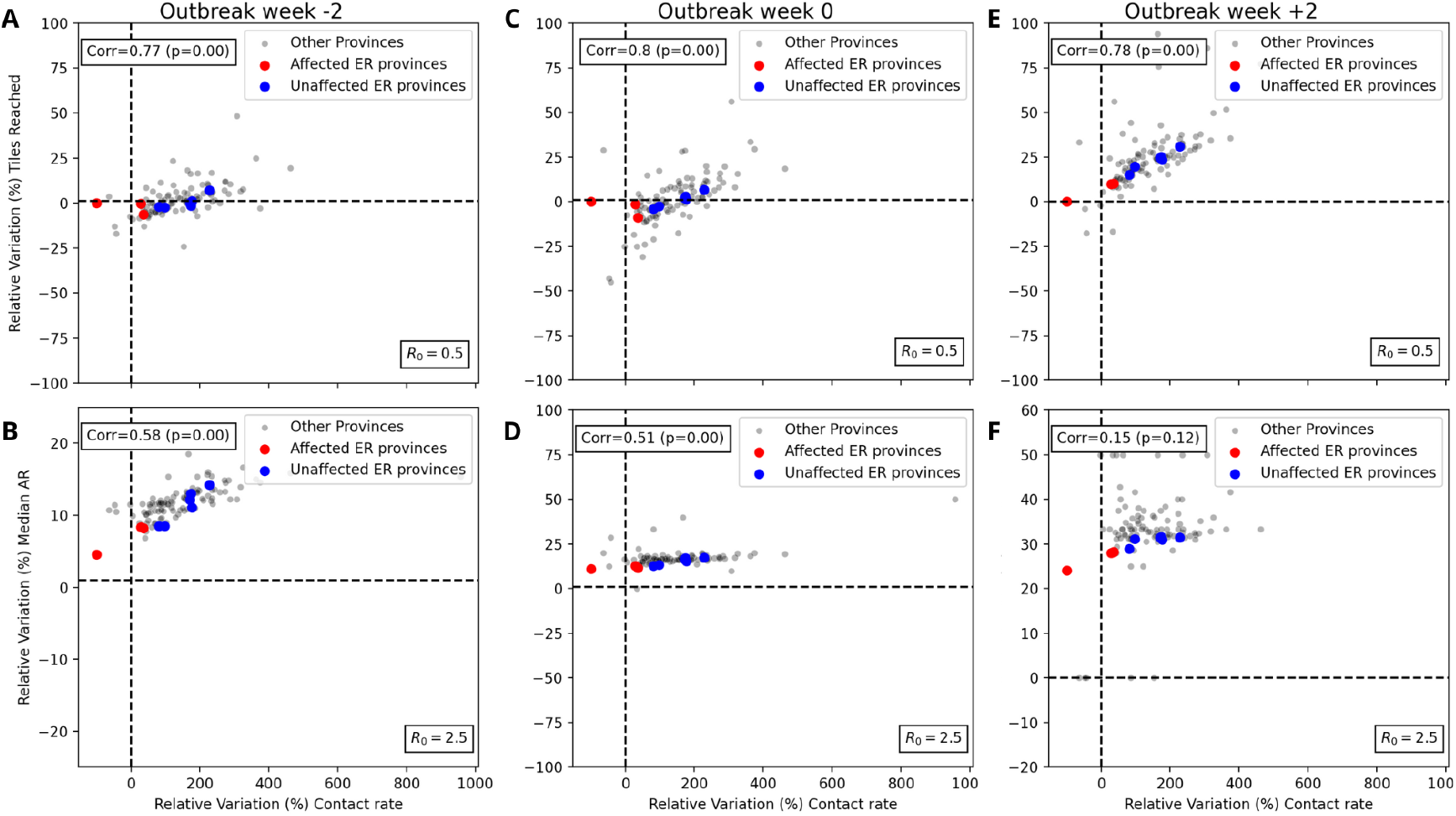
**A,C,E**. Scatterplot with the relative variation (%) of the contact rate of every province with the province of Rimini at observational week 3 and the relative variation (%) of the probability of the epidemic reaching a province with respect to the counterfactual case at 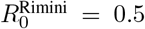 The blue dots report the affected provinces, the red dots the unaffected Emilia-Romagna provinces, the grey dots all the others. **B,D,F**. Scatterplot with the relative variation (%) of the contact rate of every province with the province of Rimini at observational week 3 and the relative variation (%) of the median outbreak size by province with respect to the counterfactual case at 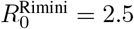.The color coding is the same as in **A,C,E**.

Here we showed a specific scenario focused on the province of Rimini and a specific time window to observe the epidemic outcome. However, we also explored other observation windows and outbreak origins in the Supplemental Material. Overall, the results matched what observed here, with some differences. Specifically, observing the epidemic outcome at 3 months instead of 4 increases the effect of the flood on the median attack rate. Observing it at 5 months instead reduces it. Nonetheless, the correlation levels in these two cases are compatible compared to what seen in Fig. 5. Moreover, the effect of the flood on an epidemic starting in Forlì-Cesena was comparable to that on an epidemic starting in Rimini. The effect of those starting in Ravenna was instead weaker.

## Discussion and Conclusion

Using close-proximity contact rates provided by Meta’s Data For Good Lab, we studied the effect of the floods in Emilia-Romagna, Italy in May 2023 on the patterns of close-proximity contacts within households, within communities and across spatial communities in Italy. The floods were associated with a marked disruption of all three types of contacts in the affected areas, whose change was in the range −40%-to-+60% in the weeks after the event, with respect to a pre-event baseline. Rimini, the Italian province most affected by the flood was also the one experiencing strongest disruptions.

Contact disruptions followed a clear pattern across affected provinces. In the immediate aftermath, contacts in households increased while contacts in the community and especially contacts with other communities decreased. This is consistent with residents being confined to their homes, and people being unable or unwilling to travel to and from the affected areas [43, 44].

The disruptions in contacts continued through several weeks after the event, but changed in sign and magnitude. Crucially, contact rates within affected communities and between affected communities and unaffected communities across Italy sharply increased, while also household contacts kept being above baseline. The increase in external contacts may be consistent with the influx of aid workers and volunteers coming in to the affected areas from all over Italy.

The recorded changes in the contact patterns are concerning in the contexts of epidemics of respiratory pathogens such as influenza and SARS-CoV-2. The rate of local contacts was known to be positively associated with the local reproduction ratio in the context of the COVID-19 pandemic [45, 46]. As such, the evidence of rising household and community contacts in the aftermath of floods may increase the chance of outbreaks by temporarily lowering the epidemic threshold [47], or facilitate epidemic circulation of ongoing outbreaks. Also, the network of contacts among spatial communities is known to affect the spatial spread of epidemics and determine whether or not large-scale diffusion occurs [21, 48, 49]. As such, the increase of contacts between residents of affected provinces and the other provinces of Italy recorded after the floods may facilitate patterns of pathogen importation and exportation and seed new outbreaks which would not otherwise occur.

To quantify the aforementioned effects, we used a spatial stochastic metapopulation model to simulate epidemics of a respiratory pathogen and test the impact of contact disruptions on epidemic circulation, compared to a counterfactual scenario in which no floods occurred. We tested basic reproduction ratios around the epidemic threshold (*R*_0_ = 1) in the range from *R*_0_ = 0.5 to *R*_0_ = 2.5. The rationale is that, if *R*_0_ ≪ 1, outbreaks are likely to be short-lived and spatially contained and, if *R*_0_ ≫ 1, outbreaks will grow fast and at large scale. The impact of floods may instead be most marked when the reproduction ratio is close to critical, as the observed contact disruptions may be enough to temporarily generate supercritical conditions leading substantially higher rates of local and global incidence as well as to bringing the epidemic to communities which would have otherwise been unaffected. Our simulations indeed showed that floods like those under study may indeed both increase epidemic size and the likelihood of long-range spread, in the context of respiratory pathogens. We found that epidemics starting in a province affected by floods increased in size as much as 20%, both inside the province where it started and elsewhere. We also observed a similar increase in size of rare, large outbreaks (those occurring once every 1,000). All these increases were most evident when epidemics started 3 weeks after the event, thus at the same time as community and external contacts spiked, and for *R*_0_ close to the epidemic threshold, confirming the intuition that floods may temporarily drive outbreaks to well above the threshold. Further evidence of this is that the floods, and the new contact patterns it caused, drove epidemics that were well in the subcritical regime (*R*_0_ = 0.5) to reaching many parts of Italy that they would not otherwise reach, including big cities. Figure 4 tells us, for example, that an outbreak starting in Rimini three weeks after the floods was 20% more likely of reaching the city of Milan than what would have happened in the absence of floods. The implication of this is clear. An epidemic that is well below threshold in Rimini (*R*_0_ = 0.5) would have a higher chance of spreading at large scale, and for longer, should it reach a high-population density and mobility hub like Milan[50, 51]. It is important to point out, however, that these effects are short-lived. They affect the spreading pattern and size for a limited amount of time; the impact of the flood on the epidemic decreases in the months following the event, as observed for wildfires [25].

Floods, a type of extreme climatic event exhacerbated by climate change, may favor the emergence and circulation of directly-transmitted respiratory pathogens. Our study shows that population-level contact data combined with mathematical model can assess this threat, quantify this risk, and, in the future, help inform public health response and preparedness. Specifically, given that contact data such as those provided by Meta are accessible in near-real time, they can be used to analyze to monitor trends during and in the aftermath of the climate emergency, and inform response accordingly. Performing epidemic scenarios through mathematical models is instead more demanding, because it requires detailed information on disease natural history and dynamics. This information may not be available, or accurate, at the time of the emergency – for instance if the pathogen is newly emerging or of an unknown variant – and this will limit the reliability of scenarios coming from mathematical models. For this reason, we investigated under which conditions scenario assessment could be performed directly from the analysis of contact data, with no need for mathematical models. We found that, for outbreaks starting with a reproduction ratio below the epidemic threshold (*R*_0_ *<* 1) the increase in risk, attributable to the floods, that the pathogen is exported to an unaffected area is directly proportional to the flood-driven increase in contact rate between that unaffected area and the affected one, in the weeks following the floods. This means that the measured variation in contacts directly measures the risk of pathogen exportation, with no need of running explicit mathematical models. As the reproduction ratio increases the correlation between direct contacts and epidemic outcome decreases, as the spreading dynamics involves routes other than direct transmission from the two communities. This means that for high *R*_0_ running the mathematical models may become necessary.

Our study has limitations. As any human mixing data, there is a problem of representativeness and penetration used to generate the contact matrices [29, 52]. The data provider performed an estimation of both for Italy, showing good results [29]. As a matter of fact, the sample represented comprises of those using the internet the most, favoring shorter time scales effects. Second, our study is retrospective and focuses on a single flood event in Italy. Furthermore, it relies on simulated epidemics and not on surveillance data from any specific epidemics. As such, it cannot be readily generalized to other floods. Further studies should attempt to analyze different floods in different places, using the available contact data, to investigate the general contact trends in this kind of events. Also, analyzing the impact of floods directly on disease incidence would be useful, albeit it would be very hard to disentangle the effect of the floods on disease incidence from other possible drivers, without resorting to mathematical models as we did. Lastly, our counterfactual scenario was built on the premise that contact patterns right after the floods were representative of the contact patterns which would have been observed in the absence of the climatic event. This is an assumption, but it is reasonable: it was already shown that the week-to-week variation in the Colocation Map is minimal if no external disruptions occur [40], plus population-level mechanisms are known to be regulated by two timescales[53]. One is the weekly timescale which is however not relevant in our case as Colocation Maps are aggregated weekly. The second is seasonality, which however unfolds at timescales much longer than our observation period of the contact data.

Our study unveils the effect that floods may have on facilitating the emergence and spread of respiratory pathogens, and is a first attempt at quantifying it in the context of a devastating flood that happened in Italy in 2023. It also shows that the analysis of autonomously collected, large-scale contact data such as those provided by Meta, combined with mathematical models, have the potential to inform preparedness and response to these ever more common compound emergencies.

## Data Availability

Meta Colocation Maps, used to analyze the contact patterns and to infer between- and within-community mixing for stochastic simulations can be requested at https://dataforgood.facebook.com/dfg/ tools/colocation-maps

## Ackwnoledgements

We acknowledge Giulia Pullano for insightful discussions. Colocation data were available thanks to Data For Good at Meta. This study was partially supported by: Horizon Europe grant SIESTA (101131957) to E.V.

