## Supplemental Material for "How floods may affect the spatial spread of respiratory pathogens: the case of Emilia-Romagna, Italy in May 2023"

### 1 Supplemental Methods

#### 1.1 Rescaling Colocation maps

Colocation Maps, by construction, are good at measuring variations in contact rates but less good at measuring the absolute number of contacts because they lack a disease-specific definition of what contacts are. So, it was necessary to apply a correction to the data. We rescaled it using Socrates data. Let $Z$ be the total outside-household number of contacts and $H$ the within-household number of contacts, as measured by [1]. We can define the mean number of contacts that a person in community $i$ experiences with people from community $j$ as $M_{ij} = \Psi C_{ij} N_j$ and its baseline value will be $\bar{M}_{ij} = \Psi \bar{C}_{ij} N_j$, where $\Psi$ is a certain scaling constant. Thus, we can distinguish

$$M_{ij}^H = \Psi^H C_{ij}^H N_j, \tag{1}$$

$$M_{ij}^Z = \Psi^Z C_{ij}^Z N_j \tag{2}$$

with $C_{ij}^H$ being the within-household contact rate matrix and $C_{ij}^Z$ is the outside-household contact rate matrix, defined in the main article. The sum over $j$ of such quantities will therefore become the mean number of contacts of people from community $i$ with everyone else. Setting

$$\begin{cases} H = \sum_j \bar{M}_{ij}^H = \bar{M}_{ii}^H, \\ Z = \sum_j \bar{M}_{ij}^Z, \end{cases} \tag{3}$$

we can quantify the constants $\Psi^H$ and $\Psi^Z$ as

$$\begin{cases} \Psi^H = \frac{H}{\bar{C}_{ii}^H N_i}, \\ \Psi^Z = \frac{Z}{\sum_j \bar{C}_{ij}^Z N_j}. \end{cases} \tag{4}$$

To model the fact that transmission along community contact is less efficient than that along household contacts we introduced a damping parameter $\omega$: [2, 3]

$$T_i = \beta \sum_j I_j (\Psi^H C_{ij}^H + \omega \Psi^Z C_{ij}^Z) = \tag{5}$$

$$= \beta H \sum_j I_j \left( \frac{1}{\bar{C}_{ii}^H N_i} C_{ij}^H + \frac{\omega Z}{H} \frac{1}{\sum_k \bar{C}_{ik}^Z N_k} C_{ij}^Z \right). \tag{6}$$

Since $\beta$ is a free parameter we can use it to absorb $H$:

$$T_i = \beta \sum_j I_j C_{ij}, \tag{7}$$

where

$$C_{ij} = \frac{1}{\bar{C}_{ii}^H N_i} C_{ij}^H + \frac{\omega Z}{H} \frac{1}{\sum_k \bar{C}_{ik}^Z N_k} C_{ij}^Z \tag{8}$$

is our overall rate matrix used in the simulations.

### 2 Supplemental Figures

#### 2.1 Spatial sensitivity analysis

We simulated outbreak scenarios alternative to those in the main article. Alongside Rimini, we considered starting the epidemic in two other provinces, that are Ravenna and Forlì-Cesena. In both cases, we performed an analogous analysis to that reported in the main article.

##### 2.1.1 Ravenna

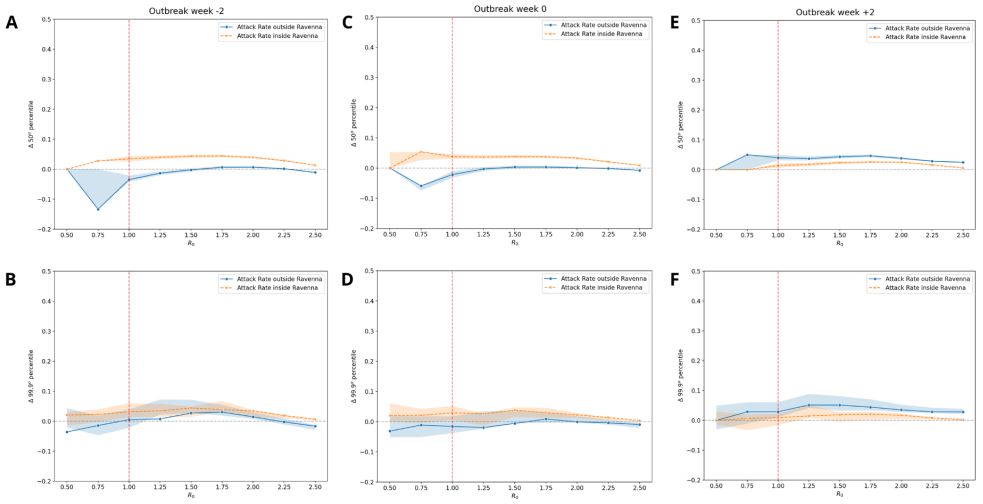

**S.1**|Relative variation with respect to the counterfactual scenario of the median (**A,C,E**) and the $99.9^{th}$ percentile value (**B,D,F**) of the Attack Rate outside and inside the province of Ravenna for different cases of outbreak week at varying values of the local reproduction ratio.In orange, the Attack Rate inside the province of Ravenna. In blue, the Attack Rate outside the province of Ravenna. The red dotted line reports the epidemic threshold ($R_0 = 1$). The shaded areas report the 95%.

First of all, we examined the impact of the flood on the size of the epidemic. Figure 1 shows the variation in median epidemic size and in the size of large epidemics, inside and outside the starting province of Ravenna, at varying basic reproductive ratio and outbreak week. Epidemic sizes inside the province only increase while outside they may also reduce according to the scenario. At varying $R_0$ the size of epidemics inside Ravenna shows always the same qualitative behavior: increasing up to $R_0 = 1.50$, then they plateau and afterwards they relax towards the x-axis. For outbreak week $-2$ and $0$, the magnitude of this variation is slightly higher then the third case. However, median size outbreaks in these two cases show a size at most $8\%$ larger than the counterfactual; for large epidemics the difference between the two cases is mostly negligible (always $< 5\%$). Moreover, the size of epidemics outside Ravenna show again a similar behavior with two exceptions. For outbreak week $-2$ and $0$, there is a decrease in the size of median outbreaks for low values of $R_0$ ($R_0 < 1$) that tends to settle towards the $0$ variation for higher values. Secondly, in case of outbreak week $+2$ for low values of $R_0$ there is an increase in the size of median outbreaks, which also proves to be more intense than inside Ravenna across all values of $R_0$. Again, also in this case the variation we see is not very intense, as at most we observe an increase of $9\%$ compared to counterfactual. In the case of large outbreaks, the behavior instead does not change.

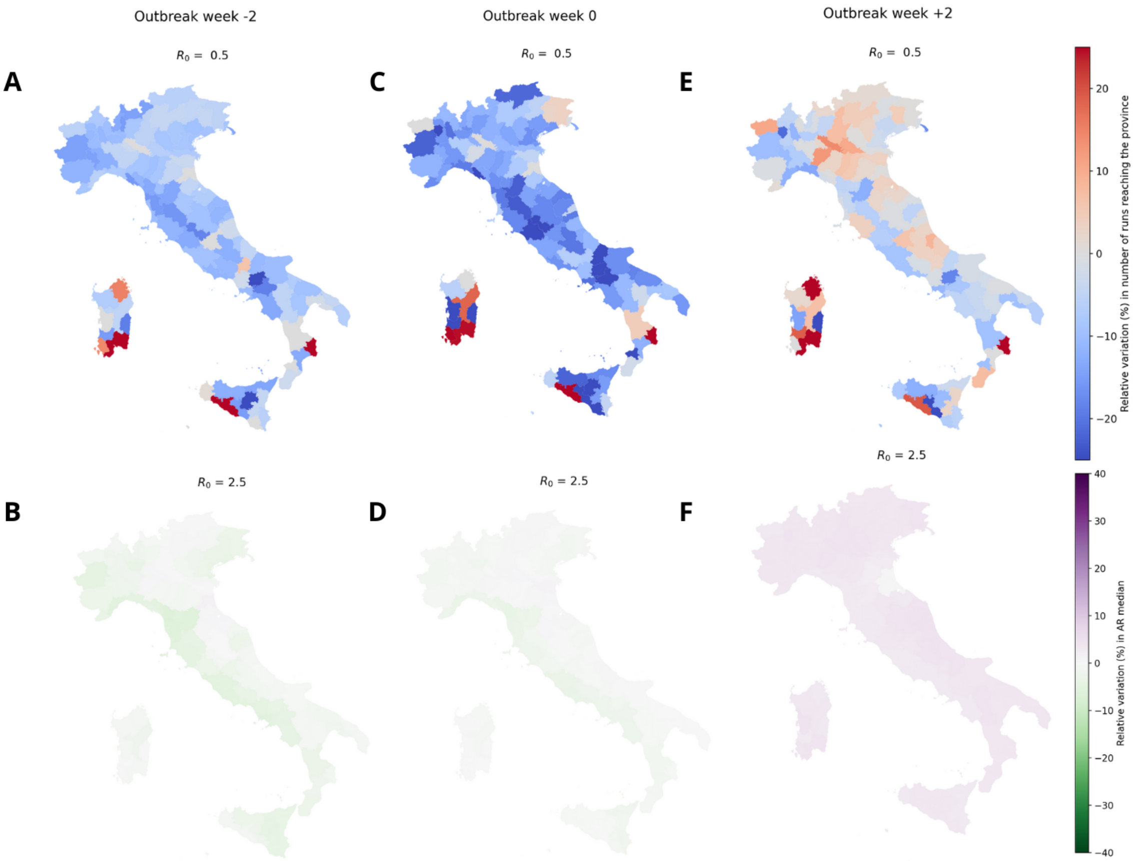

**S.2**|**A,C,E.** Administrative level 2 maps (province) of Italy, colored according to the relative variation (%) in the probability of the epidemic reaching a province, for $R_0^{\text{Ravenna}} = 0.5$, at varying outbreak week, with respect to the counterfactual case. **B,D,F.** Administrative level 2 maps (province) of Italy, colored according to the relative variation (%) in the median local Attack Rate, for $R_0^{\text{Ravenna}} = 2.5$, at varying outbreak week, with respect to the counterfactual case.

Then, we examined the impact of the flood on the probability that an epidemic well below the epidemic threshold ($R_0 = 0.5$) reaches a province (Fig. 2**A,C,E**) and the impact on the local median epidemic size while well above the threshold ($R_0 = 2.5$ - Fig. 2**B,D,F**). In the former, for outbreak weeks $-2$ and $0$, the impact is highly province-dependent, with the flood causing a few provinces to become more likely to be reached, most less likely. Overall, the effect seems to decrease in this case the probability of recording across most provinces. For outbreak week $+2$, the flood instead causes an increase in the probability to be reached in most of the provinces of Italy. For the above-threshold epidemic, outbreaks weeks $-2$ and $0$ exhibit a decrease in local outbreak size of around $5\%$ for west-coast provinces and an increase of around the same quantity for east-coast ones. For outbreak week $+2$ we observe an overall increase of the local outbreak size, with provinces seeing a moderate increase in outbreak size of up to $+10\%$.

Figure 3 investigates the correlation between the metrics of Fig. 2 and the relative change in the contact rate between each province and the province where the simulated epidemics started (Ravenna). Overall this correlation was strong and significant for the scenario below the epidemic threshold ($R_0 = 0.5$) across outbreak weeks (see also Fig. 3A,C,E), with correlation coefficient $0.76$ for outbreak week $-2$, $0.76$ for outbreak week $0$ and $0.84$ for outbreak week $+2$. In the above-threshold scenario ($R_0 = 2.5$) the correlation was markedly weaker but still significant ($pv = 0.00$) for outbreak week $-2$ (correlation $0.28$) and outbreak week $0$ (correlation $0.28$). There was instead no significant correlation in outbreak week $+2$ (correlation $0.04$, $pv = 0.66$) (see also Fig. 3B,D,F).

##### 2.1.2 Forlì-Cesena

Figure 4 shows the variation in median epidemic size and in the size of large epidemics, inside and outside the starting province of Forlì-Cesena, at varying basic reproductive ratio and outbreak week. Epidemic sizes in general only increased, regardless of the scenario. At varying $R_0$ the size of epidemics inside Forli-Cesena shows always the same qualitative behavior: increasing up to $R_0 = 1.50$, then they plateau and afterwards they relax towards the x-axis. The magnitude of this variation is unscathed by the outbreak week, showing an increase at most $10\%$ larger than the counterfactual. Moreover, the size of epidemics outside Forli-Cesena shows a similar behavior with two exceptions. For outbreak week $-2$ and $0$ (see Fig. 4**A,C**), the plateau value is reached later ($R_0 = 1.75$) and the variation settles towards a higher value than the inside case. Secondly, in case of outbreak week $+2$ (see Fig. 4**E**) for low values of $R_0$ there is an increase in the size of median outbreaks, which also proves to be more intense than inside Forlì-Cesena across all values of $R_0$. Instead the behavior in the size of larger outbreaks follows the previous trends.

We examined here the impact of the flood on the probability that an epidemic well below the epidemic threshold ($R_0 = 0.5$) reaches each province (Fig. 5**A,C,E**) and the

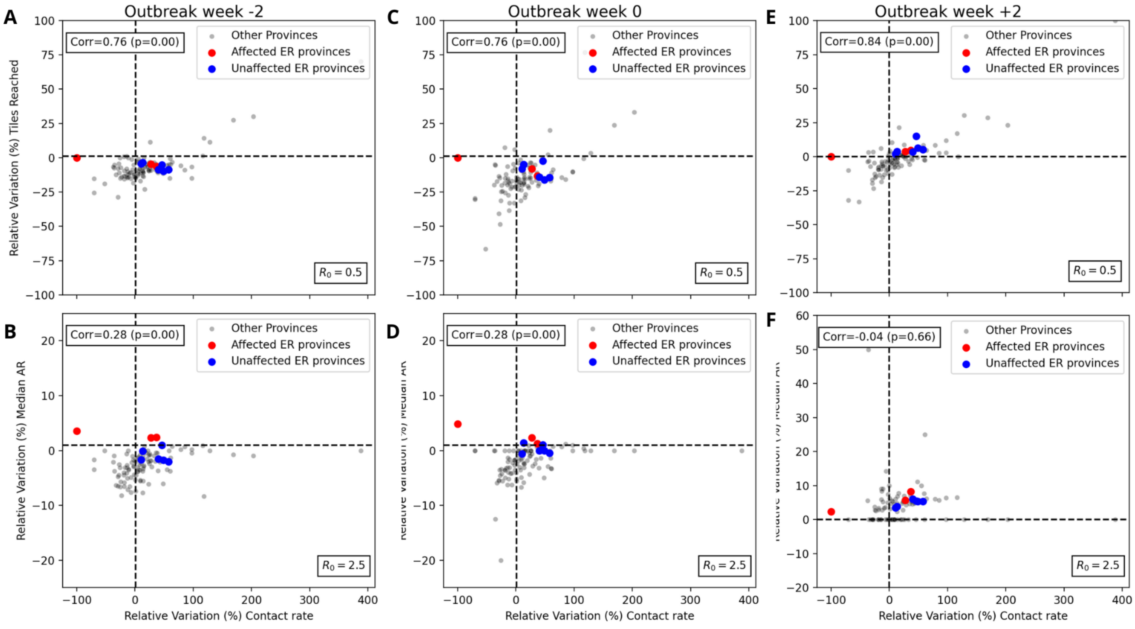

**S.3**|**A,C,E.** Scatterplot with the relative variation (%) of the contact rate of every province with the province of Ravenna at observational week 3 and the relative variation (%) of the probability of the epidemic reaching a province with respect to the counterfactual case at $R_0^{\text{Ravenna}} = 0.5$. The blue dots report the affected provinces, the red dots the unaffected Emilia-Romagna provinces, the grey dots all the others. **B,D,F.** Scatterplot with the relative variation (%) of the contact rate of every province with the province of Ravenna at observational week 3 and the relative variation (%) of the median outbreak size by province with respect to the counterfactual case at $R_0^{\text{Ravenna}} = 2.5$. The color coding is the same as in **A,C,E**.

impact on the local median epidemic size while well above the threshold ($R_0 = 2.5$ - Fig. 5**B,D,F**). In the former, for outbreak weeks $-2$ and $0$ (see Fig. 5**A,C**) the impact is highly province-dependent, with the flood causing some provinces to become more likely to be reached, other less likely. However, overall the effect seems to be stronger for nothern and southern italian provinces while less intense for center italian ones. For outbreak week $+2$ (see Fig. 5**E**), the flood instead causes an increase in the probability to be reached in most of the provinces, with only a few sparse showing a negative effect. For the above-threshold epidemic, outbreaks weeks $-2, 0$ exhibit an overall slight increase in local outbreak size of at most $10\%$. For outbreak week $+2$ the picture is homogeneous, with provinces seeing a more intense increase in outbreak size of up to $+20\%$.

Figure 6 investigates the correlation between the metrics of Fig. 5 and the relative change in the contact rate between each province and the province where the simulated epidemics started (Forlì-Cesena). Differently from the previous cases, this correlation is markedly weak for any scenario, even in the under-threshold one (see Fig. 6**A,C,E**), with correlation coefficient $0.31$ for outbreak week $-2$, $0.4$ for outbreak week $0$ and $0.41$ for outbreak week $+2$. In the above-threshold scenario ($R_0 = 2.5$) the correlation was still

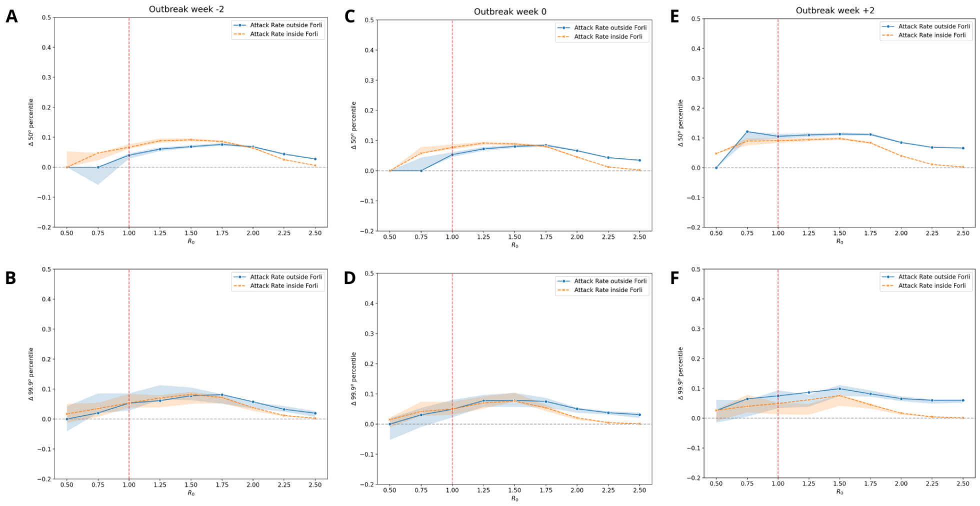

**S.4**|Relative variation with respect to the counterfactual scenario of the median (**A,C,E**) and the $99.9^{th}$ percentile value (**B,D,F**) of the Attack Rate outside and inside the province of Forli-Cesena for different cases of outbreak week at varying values of the local reproduction ratio.In orange, the Attack Rate inside the province of Forli-Cesena. In blue, the Attack Rate outside the province of Forlì-Cesena. The red dotted line reports the epidemic threshold ($R_0 = 1$). The shaded areas report the 95%.

weak though significant ($pv = 0.00$) only for outbreak week $-2$ (correlation $0.49$ - see Fig. 6**B**) . In the other two cases, there is no significant correlation (for outbreak week $0$, correlation $0.11$ and $pv = 0.25$; for outbreak week $+2$, correlation $0.13$, $pv = 0.20$ - see Fig. 6**D,F**).

#### 2.2 Temporal sensitivity analysis in Rimini

For the case of an outbreak in Rimini, we performed a similar analysis considering different weeks in which the epidemic outcome was observed.

##### 2.2.1 3 Months

Figure 7 shows the variation in median epidemic size and in the size of large epidemics, inside and outside the starting province of Rimini, at varying basic reproductive ratio and outbreak week. Epidemic sizes seem to only increase inside or outside Rimini (except for low $R_0$ in Fig. 7**B**), independently of the outbreak week. At varying $R_0$ the size of epidemics inside Rimini shows always the same qualitative behavior: increasing until a local maximum at $R_0 = 2$, afterwards it starts decreasing. Median size outbreaks (large size outbreaks) show a size at most $24\%$ $(17\%)$ larger than the counterfactual. However, the size of epidemics outside Rimini shows a rather different behavior: for any outbreak

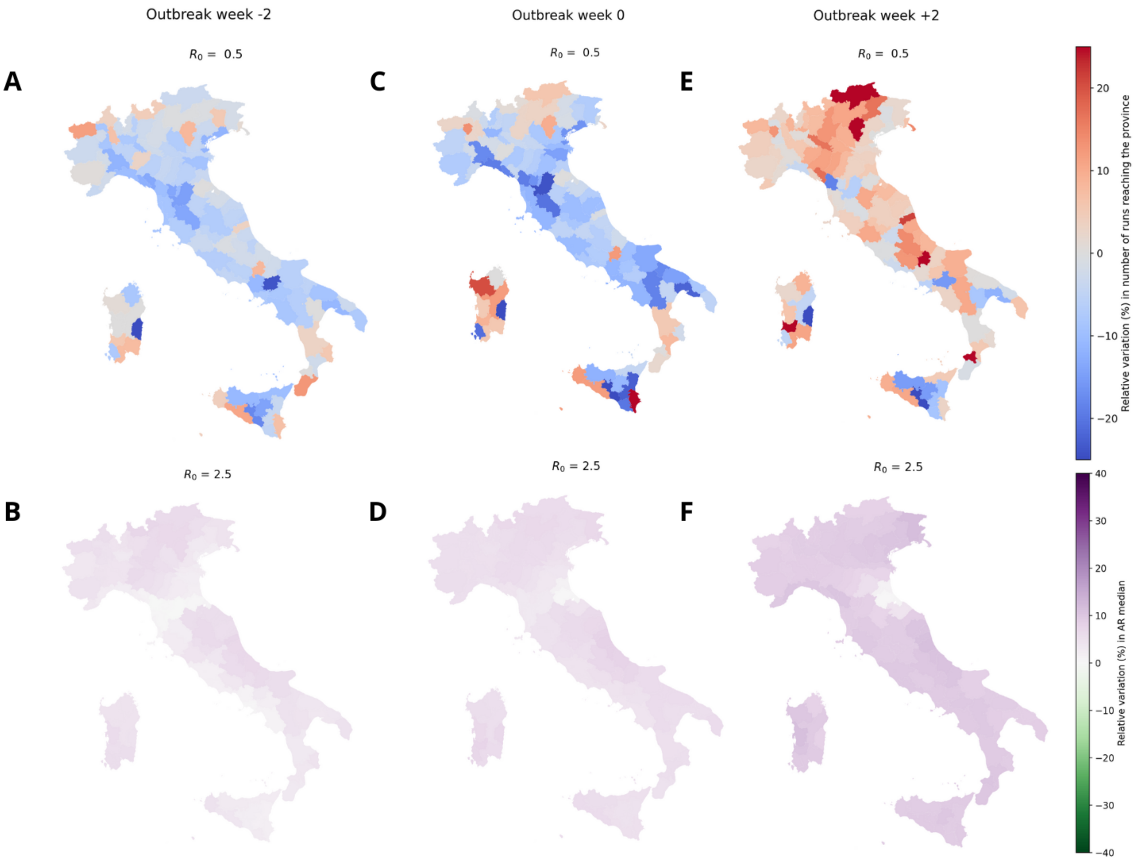

**S.5**|**A,C,E.** Administrative level 2 maps (province) of Italy, colored according to the relative variation (%) in the probability of recording an infected in a province for $R_0^{\text{Forli}} = 0.5$ per outbreak week with respect to the counterfactual case. **B,D,F.** Administrative level 2 maps (province) of Italy, colored according to the relative variation (%) in the median local Attack Rate for $R_0^{\text{Forli}} = 2.5$ per outbreak week with respect to the counterfactual case.

week and any outbreak size, it steadily increases for all $R_0$ values with two exceptions. In Fig. 7**A** and **E** for low $R_0$ there is a sudden flood-attributable increase, that disrupts the growth flow of the metric. Median size outbreaks (large size outbreaks) show a size even $50\%$ ($20\%$) larger than counterfactual (see Fig. 7**E**).

Figure 8 shows the impact of the flood on the probability that an epidemic well below the epidemic threshold ($R_0 = 0.5$) reaches each province (see Fig. 8**A,C,E**) and the impact on the local median epidemic size while well above the threshold ($R_0 = 2.5$ - see Fig. 8**B,D,F**). In the former, for outbreak weeks $-2, 0$ (see Fig. 8**A,C**) the impact is highly province-dependent, with the flood causing some provinces to become more likely to be reached, other less likely. For outbreak week $+2$ (see Fig. 8**E**), the flood instead causes an increase in the metric in almost every province of Italy, with a few exceptions in the south. For the above-threshold epidemic, the median Attack Rate shows a positive increase for every province and every outbreak scenario with one exception. While for

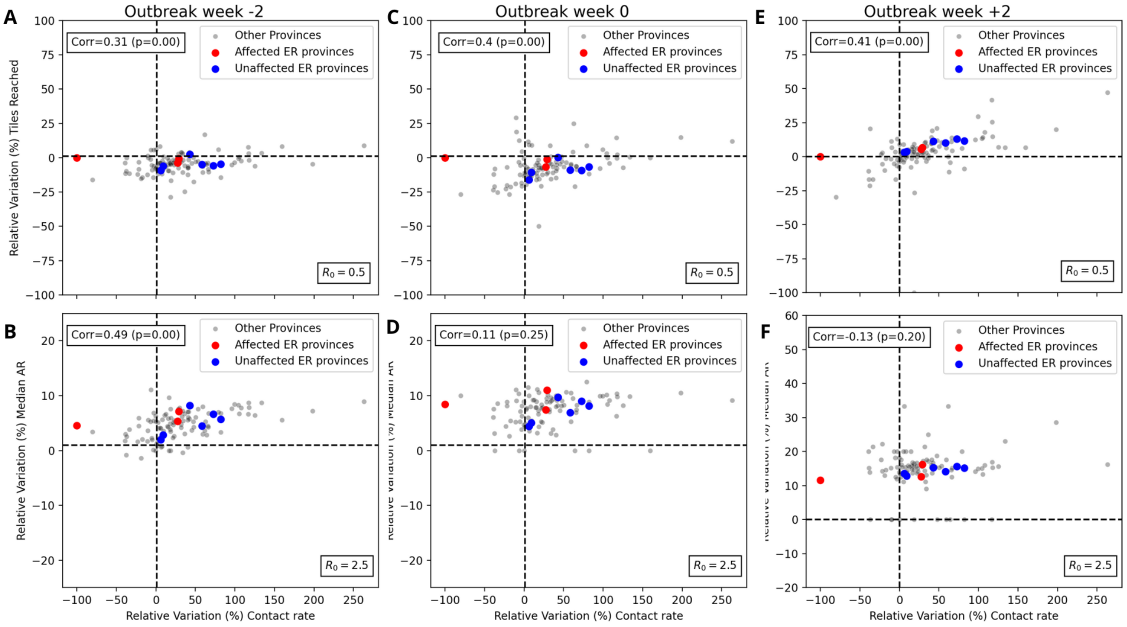

**S.6**|**A,C,E.** Scatterplot with the relative variation (%) of the contact rate of every province with the province of Forlì-Cesena at observational week 3 and the relative variation (%) of the probability of the epidemic reaching a province with respect to the counterfactual case at $R_0^{\text{Forli}} = 0.5$. The blue dots report the affected provinces, the red dots the unaffected Emilia-Romagna provinces, the grey dots all the others. **B,D,F.** Scatterplot with the relative variation (%) of the contact rate of every province with the province of Forlì-Cesena at observational week 3 and the relative variation (%) of the median outbreak size by province with respect to the counterfactual case at $R_0^{\text{Forli}} = 2.5$. The color coding is the same as in **A,C,E**.

outbreak week $-2$ and $0$ (see Fig. 8**B,D**), the map is homogeneous with local positive spikes in some provinces, for outbreak week $+2$ (see Fig. 8**F**) the effect becomes more heterogeneous according to the province with a handful of provinces across the country showing indeed a negative effect. These effects, especially those obtained for the Attack Rate can be explained by observing that at this stage the epidemic has likely not completed its full dynamic and evolution, thus we are observing effects still in a phase of transition. Indeed, the results shown in Fig. 7 report a curve which especially outside the starting province is still evolving and it has not reached its plateau value yet.

Figure 9 investigates the correlation between the metrics of Fig. 8 and the relative change in the contact rate between each province and the province where the simulated epidemics started (Rimini). This plot shows similar results to what observed in the main part. We report a markedly strong correlation in the below-threshold scenario ($R_0 = 0.5$, see Fig. 9**A,C,E**) with correlation coefficient $0.77$ for outbreak week $-2$, $0.78$ for outbreak week $0$ and $0.81$ for outbreak week $+2$ ($pv = 0.00$ in all cases). Instead, in the above-threshold scenario ($R_0 = 2.5$), we still observe a weak level of correlation though significant only for Fig. 9**B** (correlation $0.35$, $pv = 0.00$) while a rather not significant

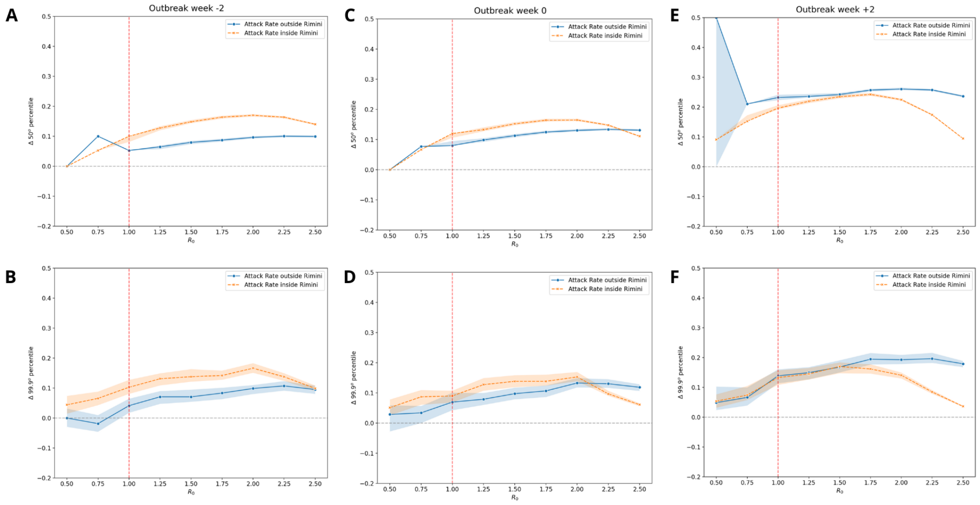

**S.7**|Relative variation with respect to the counterfactual scenario of the median (**A,C,E**) and the $99.9^{th}$ percentile value (**B,D,F**) of the Attack Rate outside and inside the province of Rimini for different cases of outbreak week at varying values of the local reproduction ratio.In orange, the Attack Rate inside the province of Rimini. In blue, the Attack Rate outside the province of Rimini. The red dotted line reports the epidemic threshold ($R_0 = 1$). The shaded areas report the 95%

correlation in the other two cases (for Fig. 9**D**, correlation $0.06$ and $pv = 0.58$; for Fig. 9**F**, correlation $0.2$, $pv = 0.17$).

##### 2.2.2 5 Months

Figure 10 shows the variation in median epidemic size and in the size of large epidemics, inside and outside the starting province of Rimini, at varying basic reproductive ratio and outbreak week. Analogously to what happens in the main part, here the epidemic sizes either increased or remained the same with respect to the counterfactual. At varying $R_0$ the variation in size of epidemics inside Rimini shows always the same qualitative behavior: increasing until a local maximum at $R_0 = 1.0 - 1.25$, then it stayed constant until $R_0 = 1.50$, afterwards it starts decreasing towards zero. For outbreak week $+2$, the median epidemic size was, at $R_0 = 1.25$, $20\%$ larger than in the counterfactual; large epidemics were $\approx 10\%$ larger (see Fig. 10**E**). The size of the epidemics outside the starting province of Rimini followed a similar trend, with two exceptions, analogous to the main result: the flood-attributable effects for large $R_0$ in outbreak week $-2$ and $0$ (see Fig. 10**A,B,C,D**) and for low $R_0$ in outbreak week $+2$ (see Fig. 10**E**)

Figure 11 examines here the impact of the flood on the probability that an epidemic well below the epidemic threshold ($R_0 = 0.5$) reaches each province (Fig. 11**A,C,E**) and

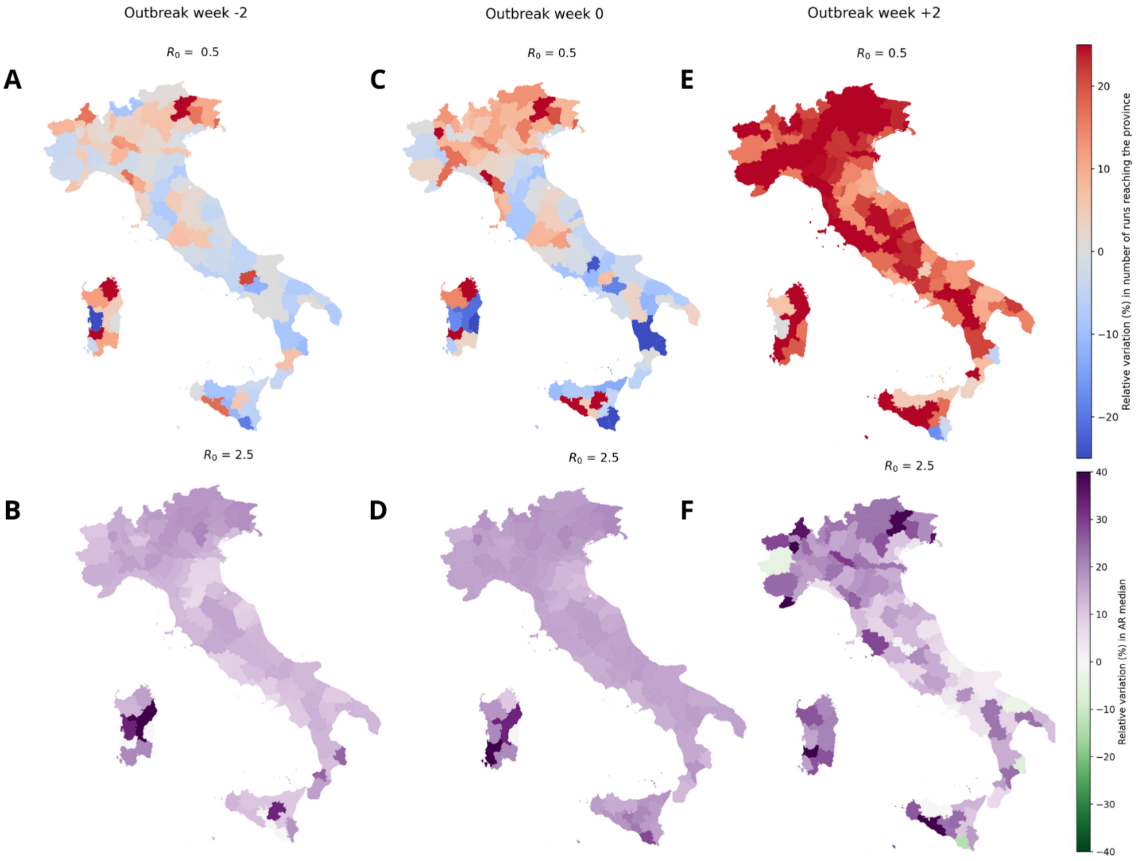

**S.8|A,C,E.** Administrative level 2 maps (province) of Italy, colored according to the relative variation (%) in the probability of the epidemic reaching a province, for $R_0^{\text{Rimini}} = 0.5$, at varying outbreak week, with respect to the counterfactual case. **B,D,F.** Administrative level 2 maps (province) of Italy, colored according to the relative variation (%) in the median local Attack Rate, for $R_0^{\text{Rimini}} = 2.5$, at varying outbreak week, with respect to the counterfactual case. The epidemic outcome was observed 3 months after the outbreak chosen.

the impact on the local median epidemic size while well above the threshold ($R_0 = 2.5$ - Fig. 11**B,D,F**). In the former, for outbreak weeks $-2, 0$ (see Fig. 11**A,C**) the impact is highly province-dependent, with the flood causing some provinces to become more likely to be reached, other less likely. Overall, the northeast provinces appear to be most positively affected while the southern ones are the most negatively affected. For outbreak week $+2$ (see Fig. 11**E**), the flood instead causes an increase in the metric in almost every province of Italy, with a few exceptions in the south. For the above-threshold epidemic, the median Attack Rate shows a generally positive increase for every province and every outbreak scenario with one exception. While for outbreak week $-2$ and $0$ (see Fig. 11**B,D**), the map is homogeneous and generally positive for almost every province (except for the affected provinces and neighbouring ones), for outbreak week $+2$ (see Fig. 11**F**) the effect becomes more heterogeneous according to the province and partic-

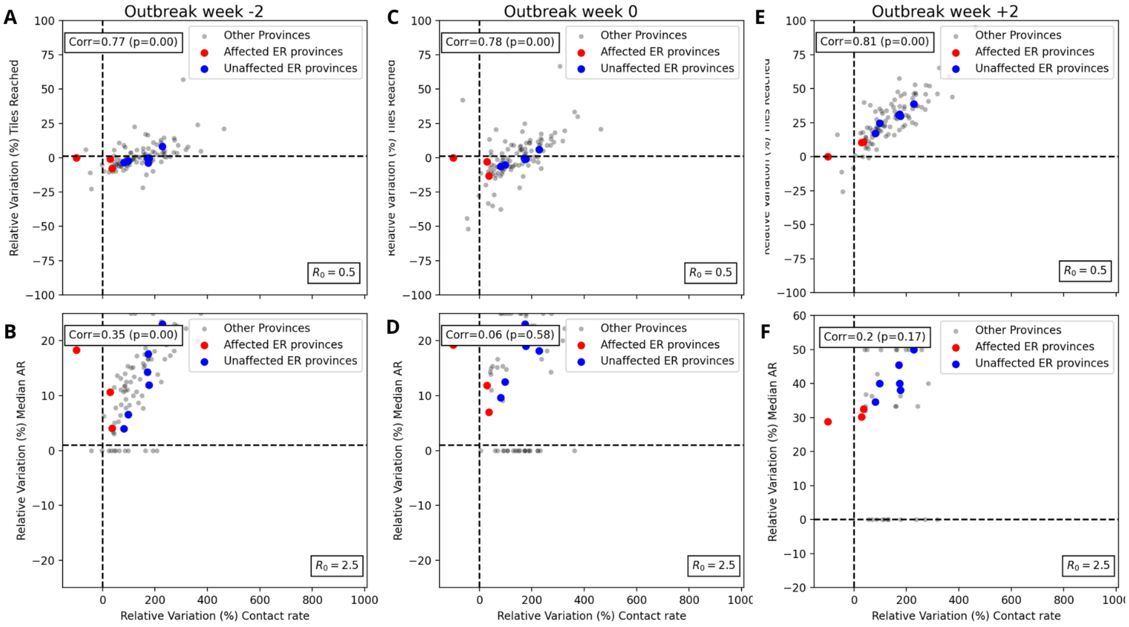

**S.9**|**A,C,E.** Scatterplot with the relative variation (%) of the contact rate of every province with the province of Rimini at observational week 3 and the relative variation (%) of the probability of the epidemic reaching a province with respect to the counterfactual case at $R_0^{\text{Rimini}} = 0.5$. The blue dots report the affected provinces, the red dots the unaffected Emilia-Romagna provinces, the grey dots all the others. **B,D,F.** Scatterplot with the relative variation (%) of the contact rate of every province with the province of Rimini at observational week 3 and the relative variation (%) of the median outbreak size by province with respect to the counterfactual case at $R_0^{\text{Rimini}} = 2.5$. The color coding is the same as in **A,C,E**. The epidemic outcome was observed 3 months after the outbreak chosen

ularly weakened in the center region of Italy, where the variation recorded is $< 5\%$. At this stage, we can expect the epidemic outcome to have well absorbed and worn out the flood effect. Thus, the real case and the counterfactual start to become much similar and reaching analogous results. This can be supported by what observed in Fig. 10, where for high $R_0$ the effect inside the province seems to be negligible compared to counterfactual and outside it is rapidly evolving towards the 0.

Figure 12 investigates the correlation between the metrics of Fig. 11 and the relative change in the contact rate between each province and the province where the simulated epidemics started (Rimini). Again, this plot shows similar results to what observed in the main part. We report a markedly strong correlation in the below-threshold scenario ($R_0 = 0.5$, see Fig. 12**A,C,E**) with correlation coefficient $0.8$ for outbreak week $-2$, $0.78$ for outbreak week $0$ and $0.75$ for outbreak week $+2$ ($pv < 0.05$ in all cases). Instead, in the above-threshold scenario ($R_0 = 2.5$), we still observe a weak level of correlation though significant, in this case, for Fig. 12**B** and **D** (the former correlation $0.52$, the latter correlation $0.32$, in both cases $pv = 0.00$). Finally, a rather not significant correlation for

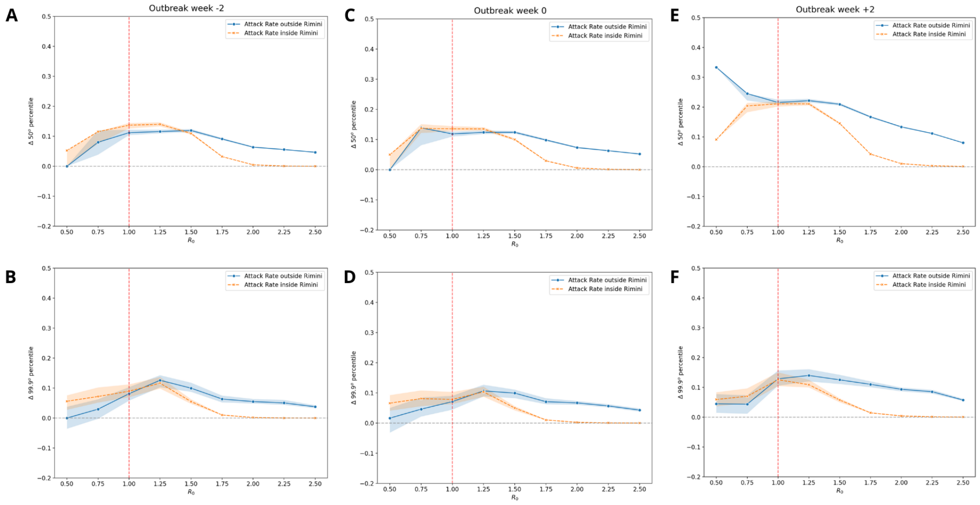

**S.10**|Relative variation with respect to the counterfactual scenario of the median (**A,C,E**) and the $99.9^{th}$ percentile value (**B,D,F**) of the Attack Rate outside and inside the province of Rimini for different cases of outbreak week at varying values of the local reproduction ratio.In orange, the Attack Rate inside the province of Rimini. In blue, the Attack Rate outside the province of Rimini. The red dotted line reports the epidemic threshold ($R_0 = 1$). The shaded areas report the 95%.

outbreak week $+2$ (correlation $0.14$ and $pv = 0.15$ - see Fig. 12**F**).

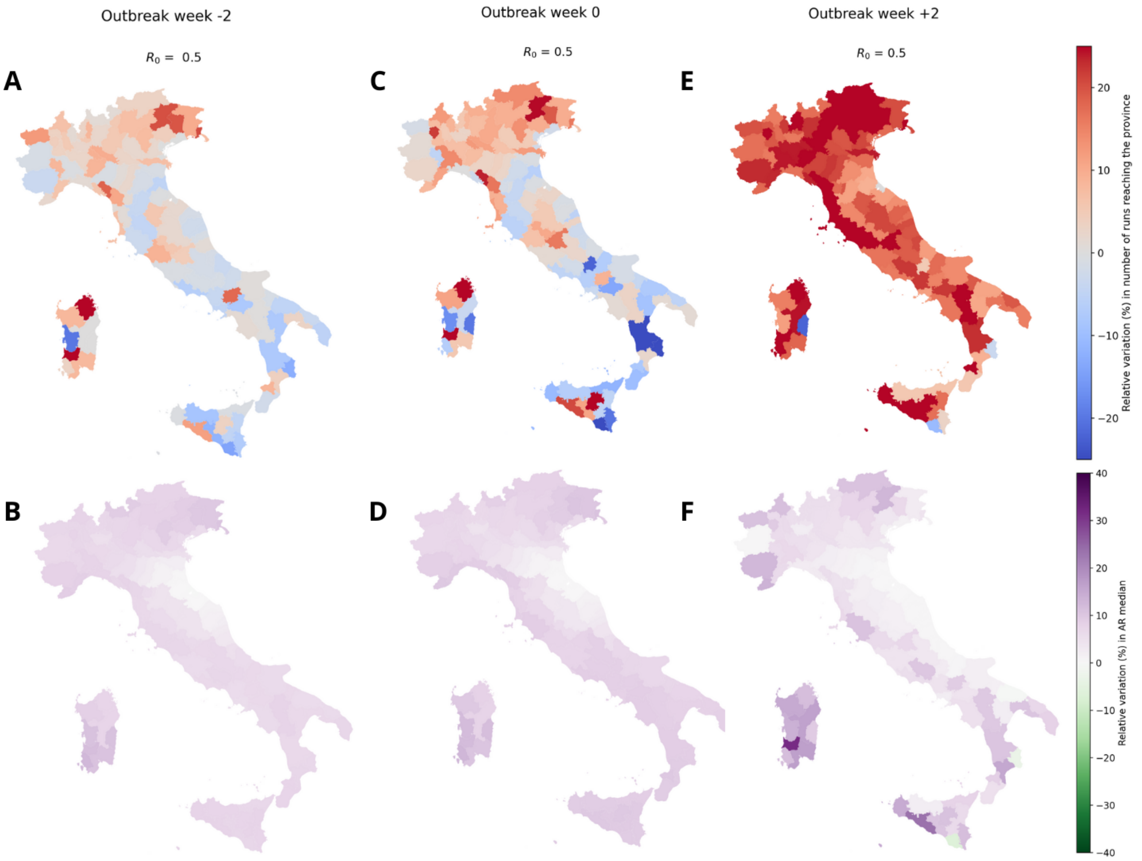

**S.11**|**A,C,E.** Administrative level 2 maps (province) of Italy, colored according to the relative variation (%) in the probability of the epidemic reaching a province, for $R_0^{\text{Rimini}} = 0.5$, at varying outbreak week, with respect to the counterfactual case. **B,D,F.** Administrative level 2 maps (province) of Italy, colored according to the relative variation (%) in the median local Attack Rate, for $R_0^{\text{Rimini}} = 2.5$, at varying outbreak week, with respect to the counterfactual case. The epidemic outcome was observed 5 months after the outbreak chosen.

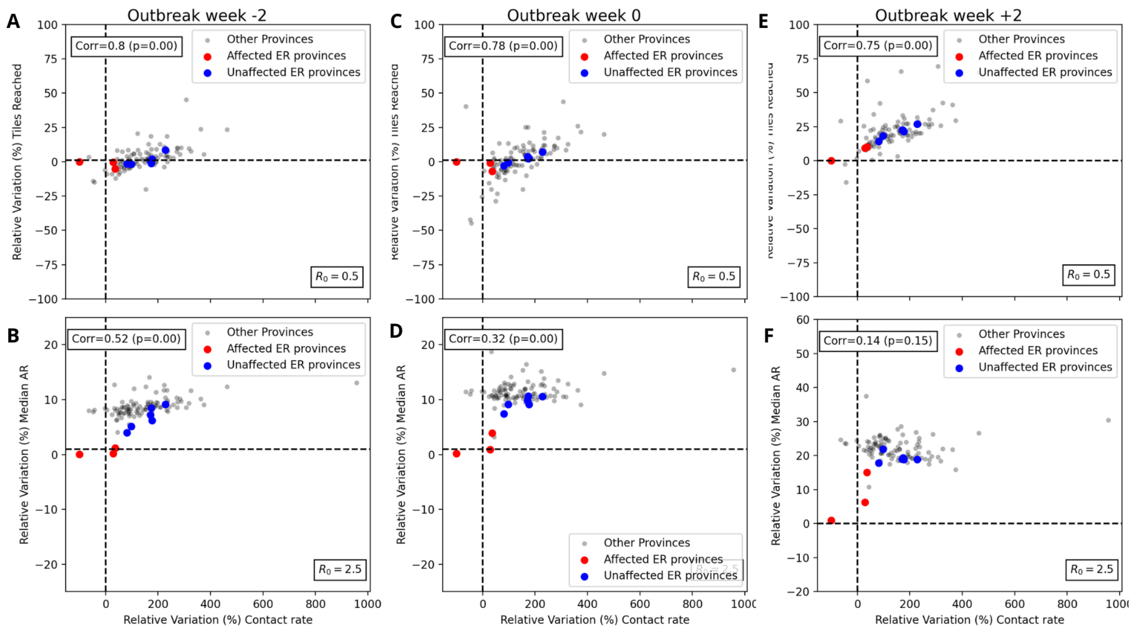

**S.12**|**A,C,E.** Scatterplot with the relative variation (%) of the contact rate of every province with the province of Rimini at observational week 3 and the relative variation (%) of the probability of the epidemic reaching a province with respect to the counterfactual case at $R_0^{\text{Rimini}} = 0.5$. The blue dots report the affected provinces, the red dots the unaffected Emilia-Romagna provinces, the grey dots all the others. **B,D,F.** Scatterplot with the relative variation (%) of the contact rate of every province with the province of Rimini at observational week 3 and the relative variation (%) of the median outbreak size by province with respect to the counterfactual case at $R_0^{\text{Rimini}} = 2.5$. The color coding is the same as in **A,C,E**. The epidemic outcome was observed 5 months after the outbreak chosen
